# Relationship between the concentration of ergothioneine in plasma and the likelihood of developing pre-eclampsia

**DOI:** 10.1101/2022.12.19.22283617

**Authors:** Louise C. Kenny, the SCOPE Consortium, Leslie W Brown, Paloma Ortea, Robin Tuytten, Douglas B. Kell

**Affiliations:** Department of Women’s and Children’s Health, Faculty of Health and Life Sciences, University of Liverpool, Liverpool L7 8TX, UK; Metabolomic Diagnostics, Cork, Ireland; Dept of Biochemistry and Systems Biology, Institute of Systems, Molecular and Integrative Biology, University of Liverpool, Crown St, Liverpool L69 7BX; The Novo Nordisk Foundation Centre for Biosustainability, Technical University of Denmark, Kemitorvet 200, 2800 Kgs Lyngby, Denmark

**Keywords:** pre-eclampsia, prevention, ergothioneine, metabolomics, nutraceutical, mushrooms

## Abstract

Ergothioneine, an antioxidant nutraceutical mainly at present derived from the dietary intake of mushrooms, has been suggested as a preventive for pre-eclampsia. We analysed early pregnancy samples for a cohort of 432 first time mothers as part of the Screening for Endpoints in Pregnancy (SCOPE, European branch) project to determine the concentration of ergothioneine in their plasma. There was a weak association between the ergothioneine levels and maternal age, but none for BMI. Of these 432 women, 97 went on to develop pre-term (23) or term (74) pre-eclampsia. If a threshold was set at the 90^th^ percentile of the reference range in the control population (≥ 462 ng/mL), only one of these 97 women (1%) developed pre-eclampsia, versus 97/432 (22.5%) whose ergothioneine level was below this threshold. One possible interpretation of these findings, consistent with previous experiments in a reduced uterine perfusion model in rats, is that ergothioneine may indeed prove protective against pre-eclampsia in humans. An intervention study of some kind now seems warranted.

## 1. Introduction

Pre-eclampsia (PE) is a multi-system disorder of pregnancy, characterized by gestational hypertension and the new-onset of proteinuria and/or another maternal organ dysfunction [1-3]. It is widely considered to develop in two stages [4] [5]: an initial poor placentation [6] followed by oxidative stress and inflammation [7, 8]. As with many other chronic, inflammatory diseases [9], there is also substantial evidence for the involvement of a microbial component [10, 11]. PE can affect 3-5% of pregnancies worldwide [12], and has the potential to be life-threatening; the only real clinical recourse currently available the premature delivery of the fetus. Early markers for predicting pre-eclampsia are thus highly desirable.

Although some soluble protein markers such as sFlt1 and PlGF have proven to be of value (e.g. [13, 14]), a number of studies have developed the idea that small molecules (i.e. metabolomics [15, 16]) have the potential to provide both diagnostic information, and, via mechanistic reasoning, potentially treatments [17-29].

Ergothioneine is an important antioxidant nutraceutical (commonly derived from the dietary intake of mushrooms) [30-34], for which humans have evolved at least one transporter [35, 36]. In one large scale metabolomics study [37] it was by some distance the molecule most associated with the prevention of adverse cardiovascular outcomes. Its levels have also been linked with lowered incidences of cognitive defects [38], and it prevented some of the main symptoms in the rat RUPP model of pre-eclampsia [39]. In view of the above, and its status as a potent antioxidant, it was thus considered plausible to have some utility in the prevention and/or diagnosis of PE in humans [40].

The international Screening for Pregnancy Endpoints (SCOPE) study [25, 41-43] (http://scopestudy.net) bio-banked early pregnancy blood samples from a large cohort of first time pregnant women, 4.9% whom developed pre-eclampsia as defined [44] by the International Society of the Study of Hypertension in Pregnancy [41]. Pre-term pre-eclampsia was defined as disease necessitating delivery before 27 weeks’ gestation. The SCOPE study therefore provided the opportunity to assess whether there was any relation between their levels of ergothioneine and the likelihood of developing early or late-onset pre-eclampsia. The present study reports on a secondary analysis of ergothioneine levels in a previously reported case-control study assessing a panel of metabolite biomarker candidates for pre-eclampsia risk assessment at 15 +/-1 week of gestation [25]. Women with the highest level of ergothioneine had a significantly reduced likelihood of developing either preterm or term pre-eclampsia. One interpretation of such data could be that dietary supplementation with ergothioneine in pregnancy may reduce pre-eclampsia risk, and this is obviously now worth testing.

## 2. Results

### 2.1. Overall summary of the cohort studied

The nested Case-Control study was defined in the SCOPE – Europe cohort as described fully in [25], and the demographics (Table 1 of [25]) are not repeated here. For this work, we leveraged calibration data available in the previous study to estimate the ergothioneine blood levels (ng/mL) in 432 women of whom 335 did not develop PE, 23 suffered preterm PE, and 74 suffered from term PE.

**Table 1.**
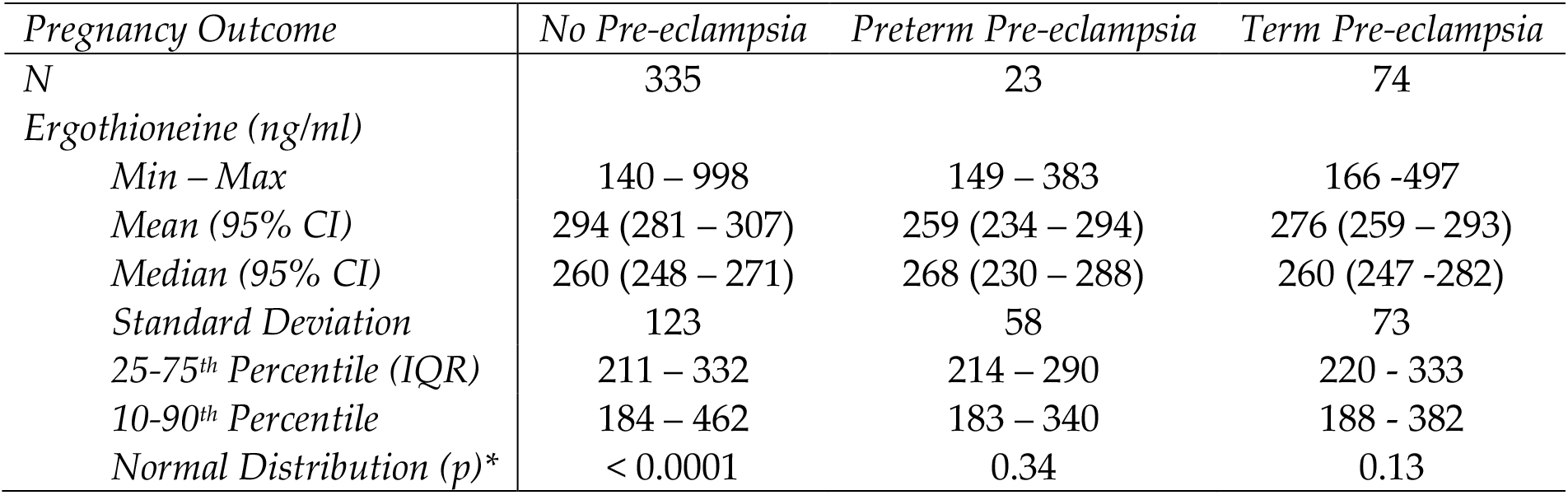
Frequentist statistical analysis of the three pregnancy outcome classes here considered; * Shapiro-Wilk test; p< 0.05: Reject Normality; IQR: Inter Quartile Range; 95% CI: 95% Confidence interval

### 2.2 Ergothioneine levels in the study population

In Figure 1 the distribution of ergothioneine levels in the study-subjects are plotted.

**Figure 1.**
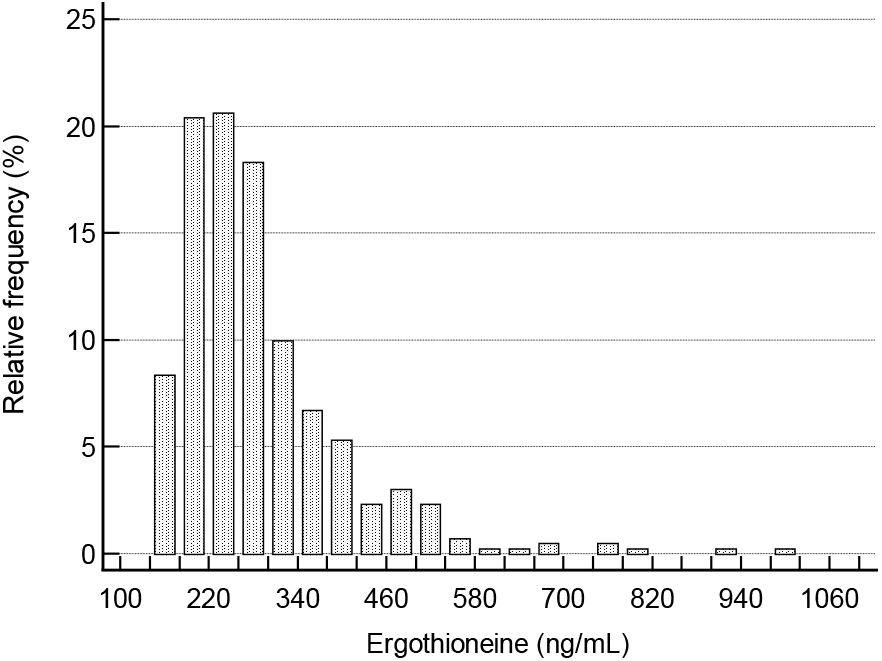
Distribution of ergothioneine levels in the study population.

The levels range from ∼140 ng/ ml to 998 ng/ml, with a median level of ∼260 ng/ml. From Figure 1, it is clear that ergothioneine levels are not normally distributed within the study population, but that the long-tailed distribution is skewed to higher concentrations.

The distribution of ages may be observed in Figure 2, where it may also be seen that there was a weak yet significant correlation of ergothioneine levels with age (r^2^= 0.08; p<0.0001). However, there was no significant relationship (r = -0.07; p=0.14) between ergothioneine levels and BMI (Figure 3).

**Figure 2.**
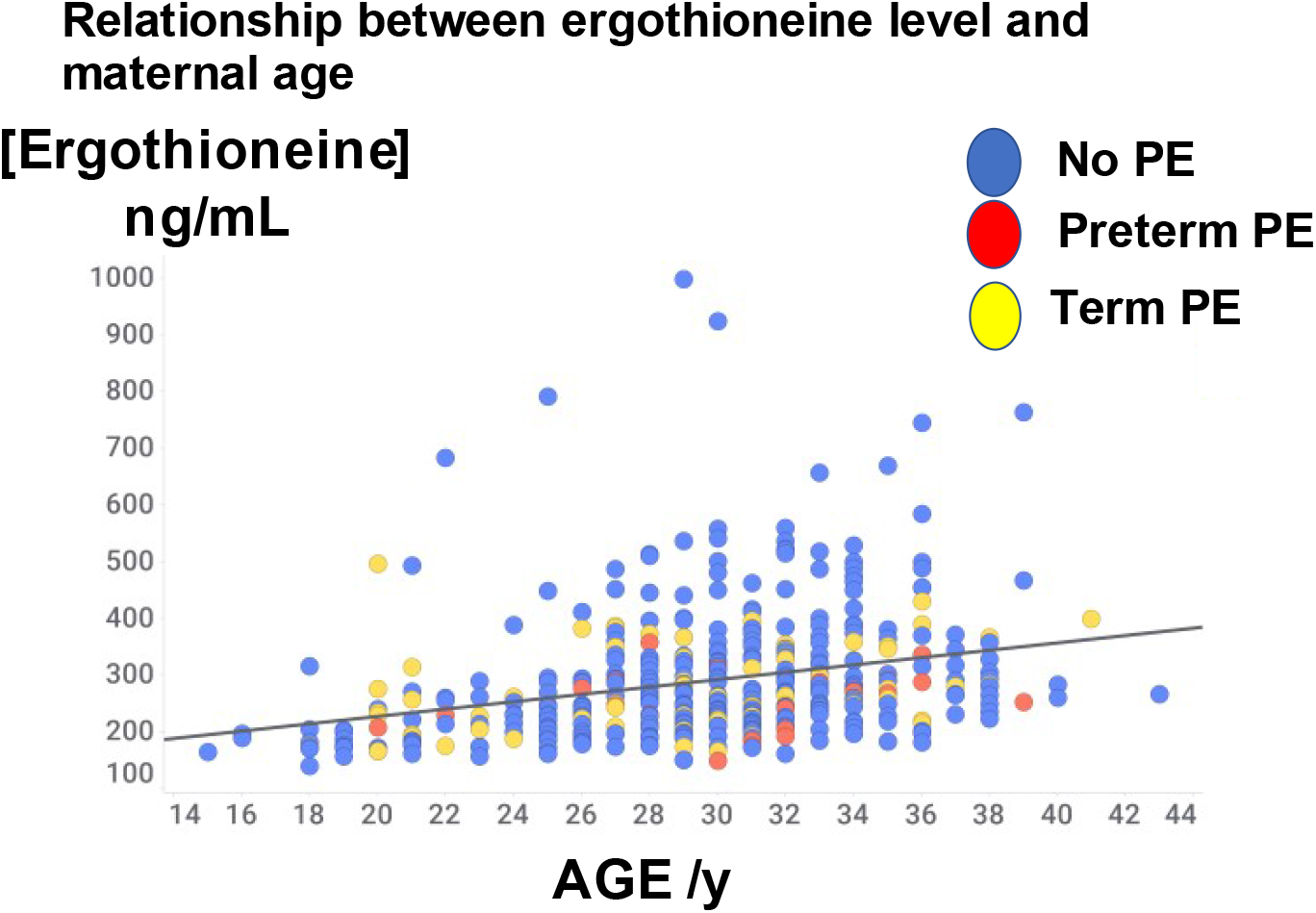
Relationship between ergothioneine level and maternal age.

**Figure 3.**
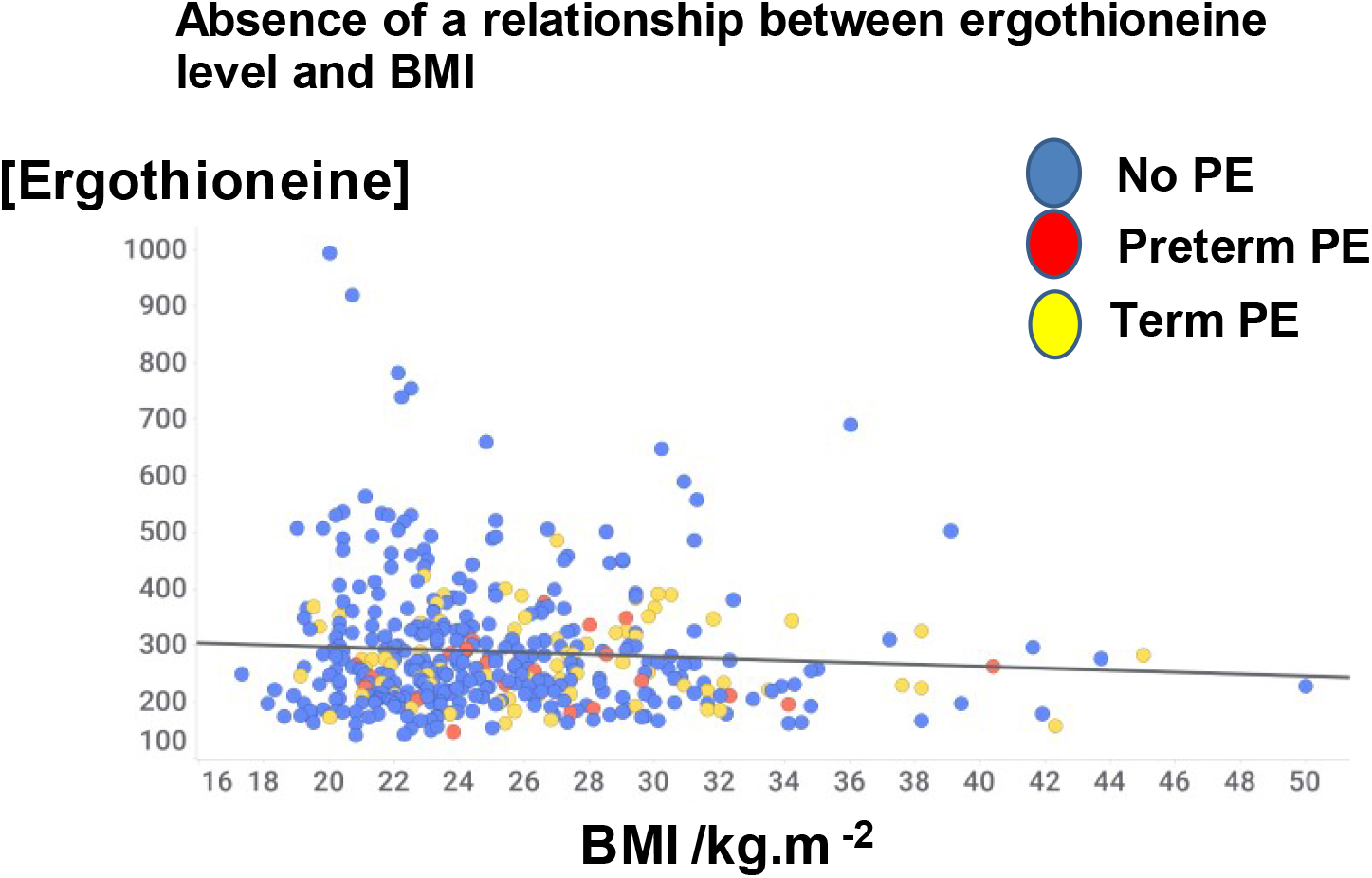
Lack of relationship between ergothioneine level and BMI. Data have been jiggered in the x-axis direction for clarity

### 2.3. Levels of ergothioneine in control and in preclamptic women

A chief finding here is illustrated in Fig 4, where we show the relationship between the levels of ergothioneine and whether the women concerned developed pre-eclampsia, whether at term or pre-term. Taking a threshold of >462 ng/mL, which is equivalent to the 90^th^ percentile in the control population, we see that only 1/74 developed PE at term, and 0/23 developed PE pre-term, i.e. only 1/97 (1%) when these numbers are combined developed any form of PE. 33/335 (10%) who did not develop PE were also above this threshold. In other words, only a single individual out of 432 examples both had a level of ergothioneine above 462 ng/mL <u>and</u> developed any form of PE.

**Figure 4.**
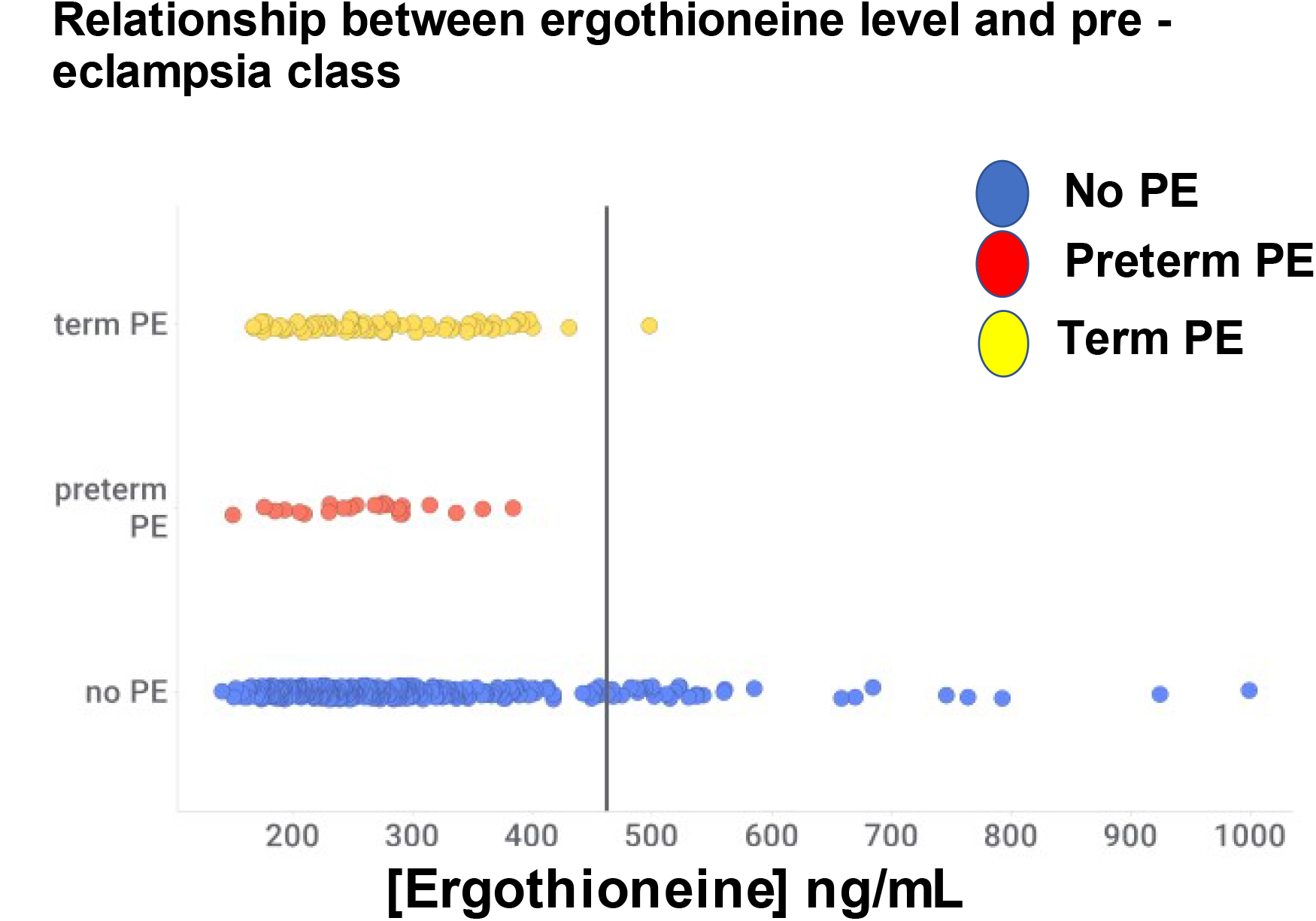
Relationship between ergothioneine level and pre-eclampsia category. Data have been jiggered in the y-axis direction for clarity

**Figure 5.**
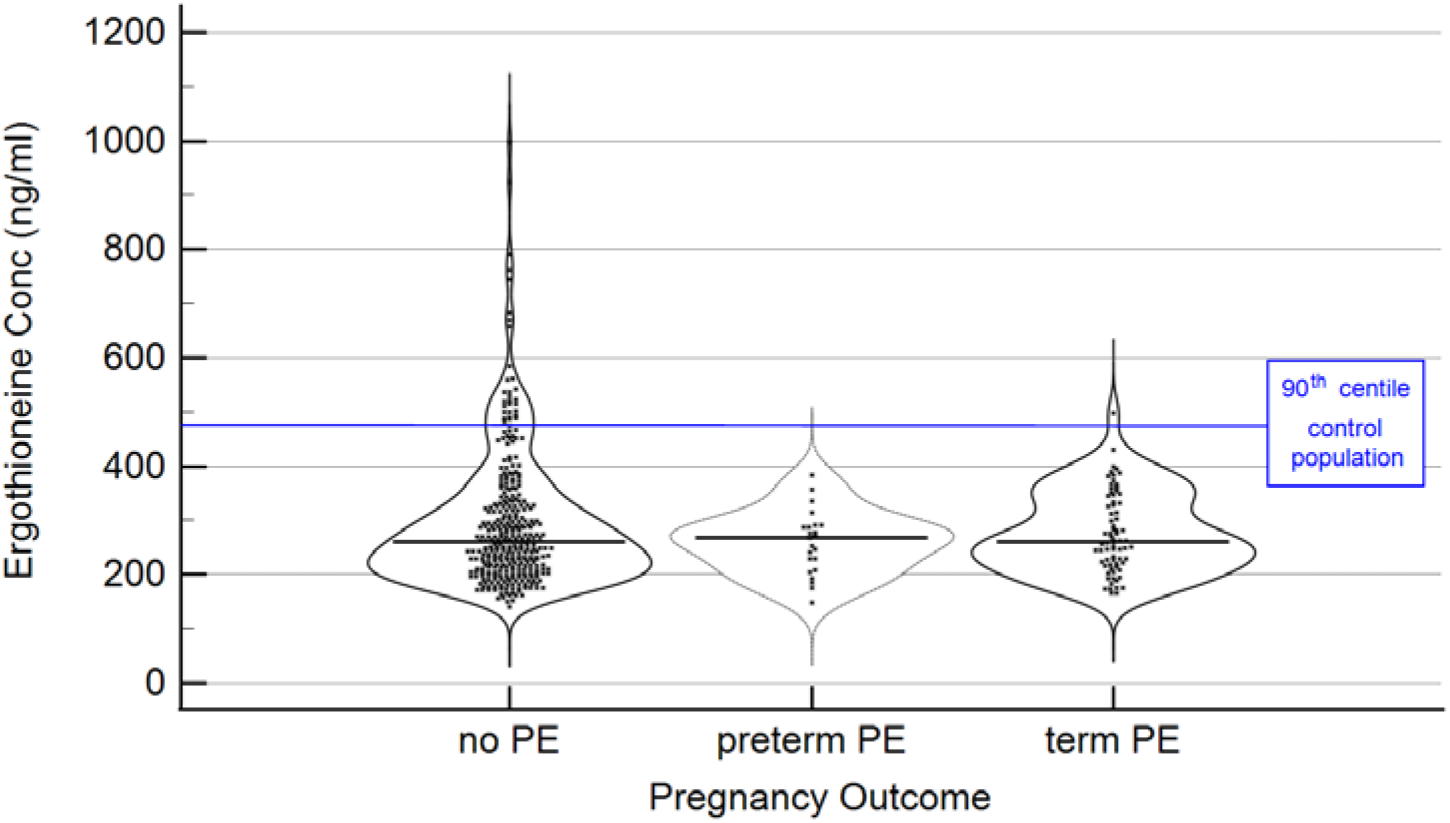
Violin plots based on individual measurements; --- median levels; blue line-90^th^ centile based on ergothioneine distribution in the control population. Data as in Figure 4 with statistical summary in Table 1.

The ergothioneine levels for those three classes are given in Table 1, where it is clear that without considering the specific distributions in detail the mean and median figures would not have indicated an association between ergothioneine and pre-eclampsia risk. From the distributions as plotted in Fig 4 it can be inferred that within the study population, there is sub-set of study participants who have markedly higher ergothioneine blood levels, giving rise to a bimodal distribution within the study population.

The relationship shown in Figure 4 (i.e. setting a cut-off at the 90^th^ centile) was post-hoc, yet it should be evident that a data-driven, machine learning analysis (to be contrasted with a frequentist statistical approach [45, 46]) would have discovered it, much as it did [22] in our first foray into pre-eclampsia metabolomics (using a low-resolution mass spectrometer that – as is still common even with high-resolution instruments [47-50] – could not identify most peaks of interest). We also note that reference-range-based rules are commonly used in clinical diagnostics to identify a population of interest, whereby patients with blood levels of a marker of interest in e.g. the top or bottom 10% of the reference interval are flagged.

Using the above 90^th^ centile as a threshold to identify a population of interest, the following Odds Ratio for developing pre-eclampsia is found in this group; OR = 0.095 with 95% Confidence Interval (0.0129 to 0.706) and significance level p<0.02.

## 3. Discussion

Stimulated by the recognition that ergothioneine might be a potentially useful therapeutic in PE [39, 40], and that it is certainly a potent (and safe [51]) antioxidant nutraceutical [30, 32-34, 52-59], we assessed its concentration in early pregnancy in a representative sample of 1^st^ time pregnant women who participated in the European branch of the SCOPE project. Certainly the range of concentrations observed was substantial, from an estimated ∼ 140 ng/ml up to 1 *μ*g/ml (Fig 3). Within the study population, the median levels were found to be non-differential between women who developed preterm-, termpre-eclampsia or women who did not develop pre-eclampsia later in pregnancy. However, within the women who had ergothioneine levels ≥ the 90^th^ percentile of the reference population, i.e., women who did not develop pre-eclampsia, only one study participant developed pre-eclampsia.

The levels of ergothioneine reflect both intake (especially from mushrooms [60, 61]) and the activity of the various ergothioneine transporters [35, 36, 62-64], and neither of these were either known or controlled. Thus, as with a related study on cardiovascular event incidence, where ergothioneine was strongly (indeed the molecule <u>most strongly</u>) associated with more favourable outcomes in terms of morbidity and mortality [37], this was a purely observational study. Ergothioneine was also the metabolite associated with the lowest hazard ratio for all-cause mortality [65], and had the third highest loading in a signature collection of healthy metabolites [66]. Mushroom consumption is also strongly associated with a lowering of all-cause mortality [67] and of the incidence of mild cognitive impairment [68]. Consequently, since certain aspects of the pre-eclampsia syndrome share hallmarks of vascular disease [69-71], this study adds weight to the idea that it might be a useful nutraceutical in the prevention of (cardio)vascular diseases more generally.

## 4. Materials and Methods

Ergothioneine was one of the compounds analysed with multiplex targeted liquid chromatography – tandem mass spectrometry assay for pre-eclampsia biomarker candidates as detailed in [25]. In the latter study, biomarker levels were expressed as relative concentrations whereby for any sample the target metabolite read-out was divided by the readout as obtained from a stable-isotope labelled metabolite internal standard spiked in equal amounts across all samples. Deuterated ergothioneine (D9) served as the stable-isotope labelled metabolite internal standard for ergothioneine quantification.

For this secondary data analysis, the relative ergothioneine concentrations were converted in estimated blood levels (ng/ml) using the calibrators co-analysed with the patient samples [25].

In brief, calibration curves were generated by means of firstly fortifying a pooled plasma (Technopath plasma with 2% K2-EDTA anticoagulant Lot PF-05171, Technopath, Ireland) with the metabolites of interest and then serially diluting the fortified sample with PBS/BSA buffer (0.01M phosphate buffer and 0.5% Albumin). The metabolites levels for the fortified matrix as well as the concentration span were estimated based on preliminary evaluations. This led to the creation of an 8-point calibration curve for all metabolites spanning a ∼20-fold dynamic range. (CAL1; relative level = 100 to CAL8, relative level = 5.83). In a separate experiment the levels of ergothioneine in the pooled plasma were determined by means of standard addition [72], yielding an estimated level of 138 ng/ml. Using this information, the ergothioneine calibration range expressed in ng/mL is easily derived (174 ng/ml (CAL8) – 752 ng/ml (CAL1)). It is noted that said calibration range covers 93.5% of all patient samples assessed; moreover, we typically find that the linear range extends beyond the set calibration range for the metabolites assessed. Verification that the ergothioneine calibrator curves effectively mitigated technical variability followed the estimation of imprecision from the analysis of 73 duplicate patient samples (independently prepared and randomly distributed across study batches) using the method of Hyslop and White [73], returning a satisfactory Coefficient of Variation (%) = 11.5%.

## Conclusions

The very striking observation that, in this cohort, only 1 individual out of 97 (1%) women with an ergothioneine level above 462 ng/ml manifested pre-eclampsia, whereas in the total cohort 97/432 (22.5%) did, demands an explanation. The easiest one is that this molecule is significantly protective towards (and may be consumed during) the development of pre-eclampsia. It is worth noting that, as well as its occurrence in all known culinary mushrooms, ergothioneine is an available nutritional supplement, whose safety has been well established [51, 74-77]. Such an analysis will best be done via a randomized controlled trial.

## Data Availability

Relevant data will be supplied as supplementary information following peer review post this preprint.

## Author Contributions

Conceptualization, DK, LK and RT; analytical methodology, LB, PO & RT; formal analysis, DK, RT.; investigation, LB, PO, RT, DK; resources; data curation, LK, PO & RT.; writing—original draft preparation, DK, RT & LB; writing—review and editing, DK, RT & LK.; funding acquisition, LK & RT. All authors have read and agreed to the published version of the manuscript.

## Funding

This study received funding from the EU-HEALTH Project IMPROvED (305169) of the Seventh Framework Programme, the goal of IMPROvED is to develop a clinically robust predictive blood test for pre-eclampsia. SCOPE was funded by the New Enterprise Research Fund, Foundation for Research Science and Technology; Health Research Council; and Evelyn Bond Fund, Auckland District Health Board Charitable Trust (New Zealand); Premier’s Science and Research Fund, South Australian Government (Australia); and Health Research Board (Ireland). The funders had no role in study design, data collection and analysis, decision to publish, or preparation of the manuscript. DBK is supported by the Novo Nordisk Foundation (grant NNF20CC0035580).

## Institutional Review Board Statement

As described [25], Ethics approval was gained from local ethics committees of each participating centre (Manchester, Leeds, and London 06/MRE01/98, Cork ECM5 (10) 05/02/08). Collection of data and biological samples complied with standardised procedures in all participating centres and was conducted in accordance with the principles of the Declaration of Helsinki.

## Informed Consent Statement

Written informed consent was obtained from all participants.

## Acknowledgments

The investigators thank the pregnant women who participated in the SCOPE study for their indispensable contribution.

## Conflicts of Interest

DBK is a named inventor on a patent (WO2020221795) describing the production of ergothioneine in yeast. Leslie W. Brown, Paloma Ortea and Robin Tuytten are employees of Metabolomic Diagnostics, and they and Louise C. Kenny are minority shareholders of Metabolomic Diagnostics. Leslie W. Brown, Robin Tuytten, and Louise C. Kenny are named inventors on a Patent Application which discloses the use of specific proteins and metabolites to predict pre-eclampsia risk, and which is assigned to Metabolomic Diagnostics, i.e., “Methods of predicting pre term birth from pre-eclampsia using metabolic and protein biomarkers”, with publication number WO2019155075A1.

## Notes

### Author Declarations

Ethics approval was gained from local ethics committees of each participating centre (Manchester, Leeds, and London 06/MRE01/98, Cork ECM5 (10) 05/02/08). Collection of data and biological samples complied with standardised procedures in all participating centres and was conducted in accordance with the principles of the Declaration of Helsinki.

